# Evaluating the utility of high-resolution proximity metrics in predicting the spread of COVID-19

**DOI:** 10.1101/2021.06.07.21258492

**Authors:** Zakaria Mehrab, Aniruddha Adiga, Madhav V. Marathe, Srinivasan Venkatramanan, Samarth Swarup

## Abstract

High resolution mobility datasets have become increasingly available in the past few years and have enabled detailed models for infectious disease spread including those for COVID-19. However, there are open questions on how such a mobility data can be used effectively within epidemic models and for which tasks they are best suited. In this paper, we extract a number of graph-based proximity metrics from high resolution cellphone trace data from X-Mode and use it to study COVID-19 epidemic spread in 50 land grant university counties in the US. We present an approach to estimate the effect of mobility on cases by fitting an ODE based model and performing multivariate linear regression to explain the estimated time varying transmissibility. We find that, while mobility plays a significant role, the contribution is heterogeneous across the counties, as exemplified by a subsequent correlation analysis. We subsequently evaluate the metrics’ utility for case surge prediction defined as a supervised classification problem, and show that the learnt model can predict surges with 95% accuracy and 87% F1-score.

## 1 Introduction

Since early 2020, the COVID-19 pandemic has severely impacted societies across the globe, making it the worst pandemic in over a century. Across the various phases, the response to this epidemic has been data-driven [1] to an extent not previously possible due, in large part, to the availability of new kinds of data and new methods of data collection and sharing. Case data have been available from the early stages of the epidemic [30, 12], enabling the use of computational models to guide policymakers. Multiple datasets ranging from clinical, genomic, to socio-behavioral aspects have been collected and shared across the public domain [38, 19].

One such dataset that has been available in an unprecedented manner, is aggregate and/or anonymized human mobility data derived from cellphone traces [7, 2]. The role of such mobility in monitoring and informing the impact of COVID-19 response efforts such as lockdowns and travel restrictions has been documented across multiple countries [5, 23, 17]. The utility of mobility data in modeling epidemic spread is intuitive, as it provides information about population movements and interactions, which affect virus transmission within a population. However, several other factors also modulate virus transmission, such as mask wearing, physical distancing, contact duration, environmental factors such as air circulation, temperature, and humidity, demographics, etc. Thus, despite successes in the use of mobility data for epidemic modeling and forecasting, it has been hard to pin down exactly how much mobility contributes to the spread of COVID-19. We have seen several scenarios, especially at universities, where a surge in interactions preceded a surge in cases, but there have been also cases where the correlation between interactions and cases is low [26].

It is important to be able to quantify the role of mobility in the epidemic since, until vaccines became available, the main tool available to policy-makers has been restricting interactions in the population that might lead to infection propagation. Being able to quantify the extent to which mobility is driving the epidemic would enable policy-makers to understand how much mobility restriction might affect cases and thus, broadly, lead to more informed decision-making.

Here we present a methodology to estimate the contribution of mobility to cases. We analyze the relationship between mobility data-derived metrics and COVID-19 case rates for fifty counties in the US corresponding to land grant universities in each state of the US; in the following we refer to them as Land Grant University Counties (LGUCs).

We proceed as follows. First we describe some related work on the use of mobility data in epidemic forecasting, especially in the context of COVID-19. We then describe the mobility data, from X-Mode [45], which we use for this study, along with the graph-based interaction metrics we derive from it. As a first task, we estimate the time-varying transmissibility for each county under study using an ODE model, and use multiple linear regression to quantify the role of mobility and the derived interaction metrics. We also report the correlations of each of the predictive variables with the case rates. Together, these tasks motivate us to study the supervised learning problem of predicting surges or peaks in cases based on the interaction metrics, at which we can get good prediction performance. We end with a discussion of limitations and extensions.

## 1.1 Summary of Results

- We use an ODE-based compartmental model to estimate a time-varying transmissibility value for the epidemic. This value encodes all the information about changes in the epidemic due to changes in mobility, human behavior, environmental conditions, and more.
- We use multivariate linear regression to estimate the contribution of proximity metrics derived from mobility data to this time-varying transmissibility as the *R*^2^ (fraction of variation explained).
- We do this analysis for fifty counties associated with major land grant universities in the US. We find that the contribution of the proximity metrics can be as high as 0.83 (for Honolulu, HI), and as low as 0.17 (for Oktibbeha, MS).
- We do a correlation analysis of the proximity metrics with the COVID-19 case data and show that different proximity metrics correlate well with the case data in different counties. Thus, while there is no single proximity metric that works well everywhere, their combination can be more predictive.
- We then show that we can use the proximity metrics to predict peaks in the case data with high reliability (high F1 score). This shows that mobility data can be used to produce actionable information for policy-making.

## 2 Related Work

Human mobility has been analyzed using multiple kinds of data, including cellphone pings [24], call detail records [15], social media geotags [47], aggregated mobility flows [42], and synthetic populations [40]. While previous research has focused on diverse topics, such as finding general patterns and laws of human mobility [18], analyzing traffic [4, 31], or studying human movements during disasters [24, 33], with COVID-19 and other epidemics, the emphasis has largely been on using mobility data to do epidemic forecasting.

The potential importance of large-scale high-resolution mobility data for COVID-19 response was realized early in the pandemic. Google released a public mobility index showing the change in mobility by region and activity type [16]. Google also shared aggregated mobility flow data with some research partners in a privacy-preserving way [42]. Similarly, Cuebiq released a mobility index and shared cellphone ping data with some research partners, also in a privacy-preserving way [11]. SafeGraph released a public data set based on anonymized cellphone pings and Points of Interest [32].

These data sets have been used in various modeling efforts, primarily focused on forecasting case rate and, in some cases, evaluating counterfactuals [7, 2]. For example, Kogan et al. [22] used datasets from multiple traces to evaluate their utility as early warning indicators for excess ILI (Influenza-like illness), confirmed cases and confirmed deaths. However, their work mainly focused on state levels whereas we focus on county levels. Moreover, they do not distinguish between the normal epidemic scenario and surges in case rates. Badr et al. [5] estimated the association between mobility data and growth of COVID cases by defining a pseudo metric for social distancing. They used an origination-destination trip dataset rather than actual pings and found strong correlation for specific timeframes but weak correlation for other timeframes. Cot et al. [10] performed a similar analysis to provide quantitative evidence of people’s compliance with social distancing after state-level mandates. They were able to use mobility data to associate the mandates with subsequent reductions in case rate growths. But these works do not explore the capability to do prediction. Warren and Skillman [43] used aggregated mobility data to observe changes in mobility due to the pandemic at both state level and county level. However, their work was purely descriptive and the dynamics of the relationship between mobility data and the case rate were not quantified. Xiong et al. [46] tried to capture this relationship through simultaneous equation model (SEM) using anonymized mobility data and found strong correlations in partially reopened regions. They also estimated an incoming second spike in the near future. However, the predictability of their equation model has not been explored. Another study by Wellenius et al. [44] analyzed the change in mobility with different state-mandated social distancing orders and studied the effectiveness of each order separately. They also attempted to quantify the relationship between time spent out-of-home and change in case rate growth. However, we take a more direct approach in the sense that our metrics are more of an intuitive indicator than time spent outside home in terms of how well people are maintaining social distancing. Wang et al. [41] took a more involved approach by employing graph neural networks (GNN) to find the association between mobility and epidemic spread. However, GNN is computationally expensive and the question of explainability still remains.

## 3 Preliminaries

### 3.1 Dataset

Our mobility data set is obtained via a third-party SDK operated by X-Mode [45]. X-Mode collaborates with various mobile applications to provide anonymous GPS traces containing persistent identifiers. Each data record is called a *ping*, which is a 4-tuple of the format *< u, x, y, t >*, where *u* is a device identifier, *x* and *y* are the longitude and latitude of the ping, respectively, and *t* is the timestamp. The data set contains ∼ 20 million unique users/day across the globe, of whom ∼10 million are in the US. There are no demographics associated with the records. The frequency of sampling the location of any user is irregular, so the number of pings associated with a device can vary substantially from day to day. The global data volume is quite large, ∼1.2TB/day. Restricted to the fifty LGUCs we consider, the data volume is ∼0.86GB/day.

In the present work, we consider fifty LGUCs, from forty-nine states. We excluded Washington County in Rhode Island (University of Rhode Island) because it had too few pings for several days within the time window we considered — Apr-Oct, 2020. However, we added the City of Charlottesville (University of Virginia) to bring the number to fifty. In our data set, the county with the fewest average unique IDs is Latah (Idaho) with 941.19 pings/day on average and the county with the most unique IDs on average is Franklin (Ohio) with 40933.28 pings/day on average.

### 3.2 Contact network

Let *P* be set of all pings *p*_*i*_ in a day where *p*_*i*_ is a 4-tuple of the format *< u*_*i*_, *x*_*i*_, *y*_*i*_, *t*_*i*_ *>*, specifying the unique device id, longitude, latitude and timestamp, respectively. We define *S* = {(*p*_*i*_, *p*_*j*_) | *p*_*i*_ ∈ *P, p*_*j*_ ∈*P, D*_*ij*_≤ *δ, T*_*ij*_ ≤ *τ* } to be the set of all pairs of pings satisfying spatial and temporal proximity. Here, *D*_*ij*_ is the distance between location of two pings *p*_*i*_ and *p*_*j*_, and *δ* is a pre-specified parameter indicating the spatial threshold. Similarly, *T*_*ij*_ is the difference between the timestamps of pings *p*_*i*_ and *p*_*j*_, and *τ* is the pre-specified temporal threshold.

Our contact network for each LGUC for each day is a dynamic network represented by an unweighted graph *G*^*t*^ (*V, E*^*t*^) with time-varying edges parameterized by *t*, where *V* ={*u*_*i*_| *p*_*i*_ ∈ *P* } is the set of all unique users (devices) and *E*^*T*^ = {(*u*_*i*_, *u*_*j*_) (*p*_*i*_, *p*_*j*_)∈ *S, min*(*t*_*i*_, *t*_*j*_) ≥ *t, max*(*t*_*i*_, *t*_*j*_) ≤ *t* + *T*_*c*_ is the set of edges between pairs of users who had an interaction between timestamp *t* and *t* + *T*_*c*_. Here, *T*_*c*_ is a predefined constant called contact time length (see Table 1) that specifies the maximum time difference between each interaction in a contact network. Using different values of t and *T*_*c*_, it is possible to obtain the contact network of interactions at different time windows and various resolutions within a given day.

**Table 1:**
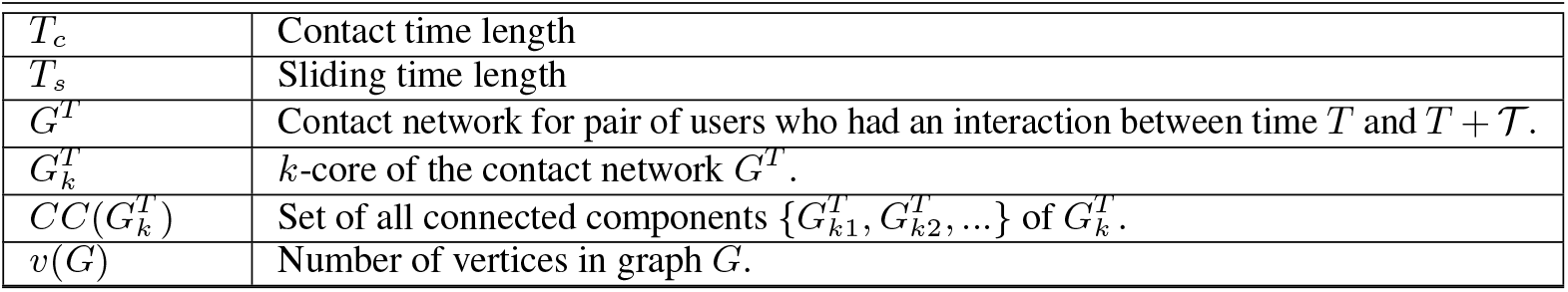
Important notations for contact network

The *k*-core of a graph is the maximal connected subgraph where all vertices of the subgraph have degree at least *k*. The *k*-core has been extensively used in social network analysis, including epidemic spread through social network [8, 21, 6] and collaborative users in the network [**?**, 48, 9, 39, 20]. We denote the *k*-core of graph *G*^*t*^ as 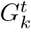, which may have multiple connected components. A connected component is a subgraph where every node is reachable from every other node. In our context, nodes in such a connected component reflect users who have interacted with each other either directly or indirectly and each user has directly interacted with at least *k* other users within the time window *t* and *t* + *T*_*c*_. Let 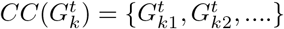 be the set of connected components in the *k*-core of *G*^*t*^. Various metrics are defined using different aggregation operations on these connected components as described in the subsequent section.

## 4 Methodology

### 4.1 Proximity metrics

We begin by extracting *stop points* from the mobility traces, using the stop point detection method in the Python scikit-mobility library [29]. The goal is to eliminate pings corresponding to situations where the device is moving, since we would like to capture those instances where a person might have an interaction with another person that is long enough for infectious disease transmission. Formally, a set of trajectory points is considered a single stop point if the trajectory points are within a minimum spatial distance from the stop point and an individual has spent a minimum duration at those trajectory points. Accordingly, the parameters for the stop point detection method are *minutes_for_a_stop*, indicating the minimum stop duration and *spatial_radius_km*, indicating the distance of the trajectory points from the stop point. The stop point is computed as the median of the trajectory points satisfying both constraints. For our experiment, we used *minutes_for_a_stop* = 5 minutes and *spatial_radius_km* = 0.01 kilometers. Once the stop points have been generated, we compute a number of metrics of interaction, described below.

#### 4.1.1 Interaction rate

We define an interaction as a co-occurrence of pings within a spatial threshold *δ* and temporal threshold *τ*. The concept of interaction is illustrated in Figure 1a. Considering a one dimensional space for clarity, the pings of three users (identified by their colors) are shown along spatial and temporal dimension. A bounding box of height *δ* and width *τ* represents the proximal regions of each ping. Based on our definition, if two bounding boxes overlap, there has been an interaction.

**Figure 1:**
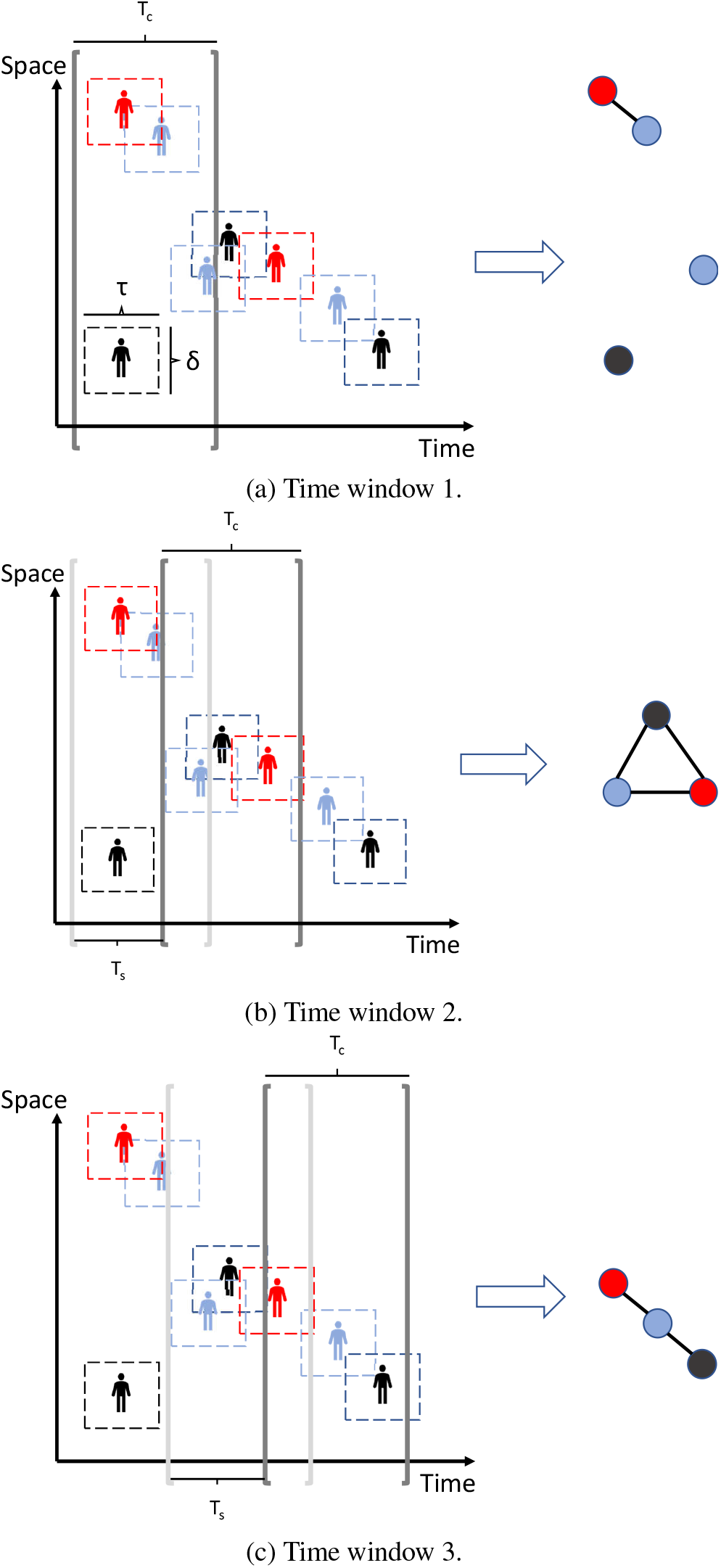
These figures illustrate the general idea of how the parameters contribute to constructing the contact networks. Bounding boxes surrounding the users denote the spatial threshold *δ* and the temporal threshold *τ*. Overlapping bounding boxes denote an interaction and there is an edge between the corresponding users in the resulting contact network (shown on the right side of each figure). Each window of length *T*_*c*_ considers a subset of pings for the current contact network. In Figure 1a, only the red and blue users at the top interact among the current set of users, whereas in Figure 1b, all the users in the set interact with each other. The time window is shifted by *T*_*s*_ for each consecutive time windows.

For each LGUC, we compute the interaction rate of the devices for every day in our study period (Apr-Oct, 2020). The interaction rate is defined as the average number of interactions of devices which participated in at least one interaction (average degree of the non-isolated nodes). For our experiments, we chose *τ* = 1 hour and *δ* = 2 meter. These values were found to give the best results in a preliminary examination.

#### 4.1.2 Graph Metrics

The extent to which a contagion spreads in a population depends significantly on the structure of the interaction graph of the population. For example, a given interaction rate *r* may be achieved by a population consisting of isolated groups of size *r* and also by an *r*-regular connected graph. In the former case, a contagion starting at one node would be restricted to at most the *r* – 1 neighbors of the node, while in the latter case it could potentially spread throughout the population. With this motivation, we generate a number of graph-based metrics (see Table 2) based on the connected components of the *k*-cores of a sequence of contact networks 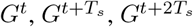 etc. Here, *T*_*s*_ is another constant we define (see Table 1) that controls the difference between two adjacent contact networks..

**Table 2:**
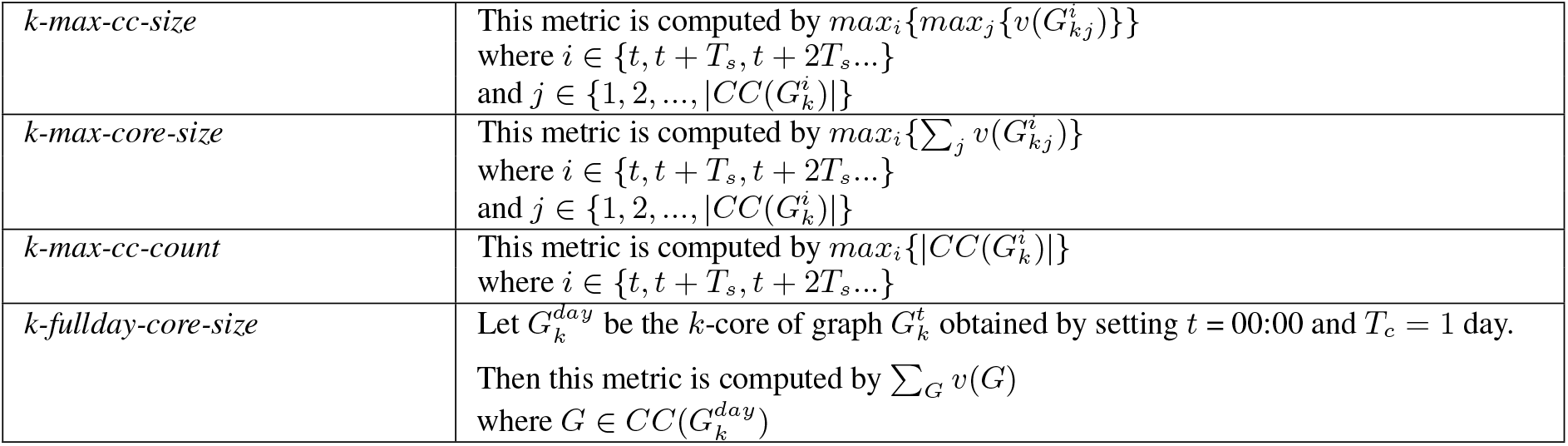
Inferred graph metrics.

The first three metrics are similar in how they are computed. Essentially, the components in the *k*-cores represent different groups or gatherings during the corresponding time window. *k-max-cc-size* captures the largest group size among these different groups whereas the *k-max-core-size* captures the total size of these groups. Both metrics are useful, as either many gatherings of small or moderate sizes or one single large gathering can be responsible for spreading the virus. The *k-max-cc-count* captures the number of such gatherings which is also necessary to understand how different communities are complying with imposed restrictions.

First, we show in Figure 1, we show how the contact network is formed with different time windows from the interactions. In the first window (Figure 1a), there are four visible pings. However, only the red user and blue user on the top have an overlapping bounding box, denoting that this is an interaction. This generates only a single edge in the resulting contact window. In Figure 1b, the window is slided by *T*_*s*_ and now all the users in this window have overlapping bounding box with each other, resulting in a clique as the contact network. In Figure 1c, the red user from the previous time window is still visible. However, now there is an overlapping boxes with a different set of pings, resulting in a different contact network than before.

Next, Using Figure 2a and Figure 2b, we explain how the first three graph metrics are generated with a more involved conceptual graph. Figure 2a shows the conceptual graph of all interactions throughout the day. An edge between two nodes in this graph means that the users corresponding to the nodes were close in space and time and the label along the edge shows the timestamps of each of the pings. For brevity, we consider that all the timestamps fall between 08:00 to 09:30. We also consider that there is no multi-edges, although this is not the actual scenario because same pair of users can interact at different times.

**Figure 2:**
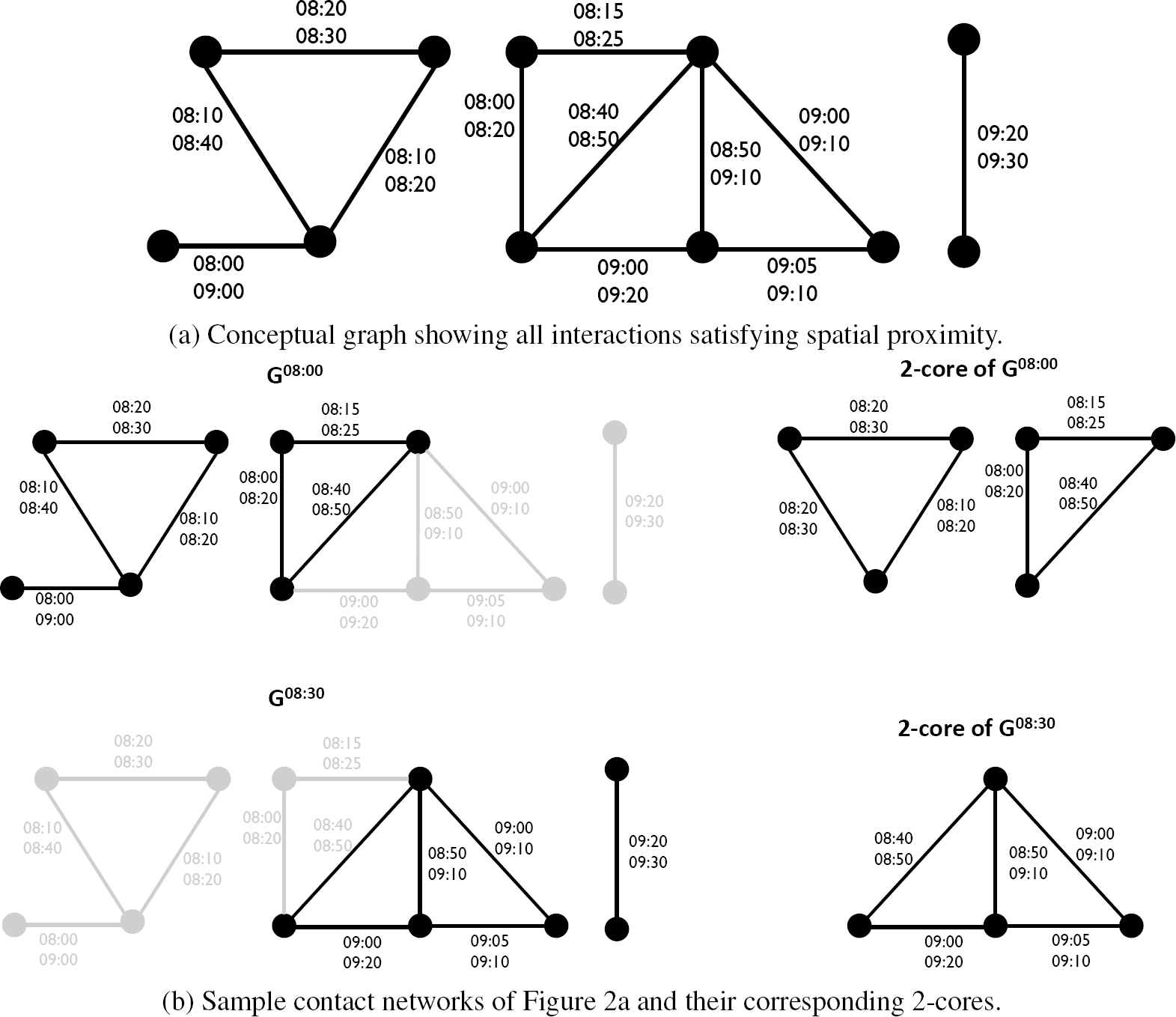
These figures illustrate the general idea of contact networks in different time windows and their *k*-cores. The time window parameters are: *T*_*c*_ = 1 hour and *T*_*s*_ = 30 minutes. Figure 2a shows the view of the contact network if we considered all the interactions at once. Figure 2b shows the contact networks for *t* = 08:00 and *t* = 08:30 and their corresponding 2-cores. See Table 2 for formal definitions of the graph metrics inferred from these *k*-cores.

**Figure 3:**
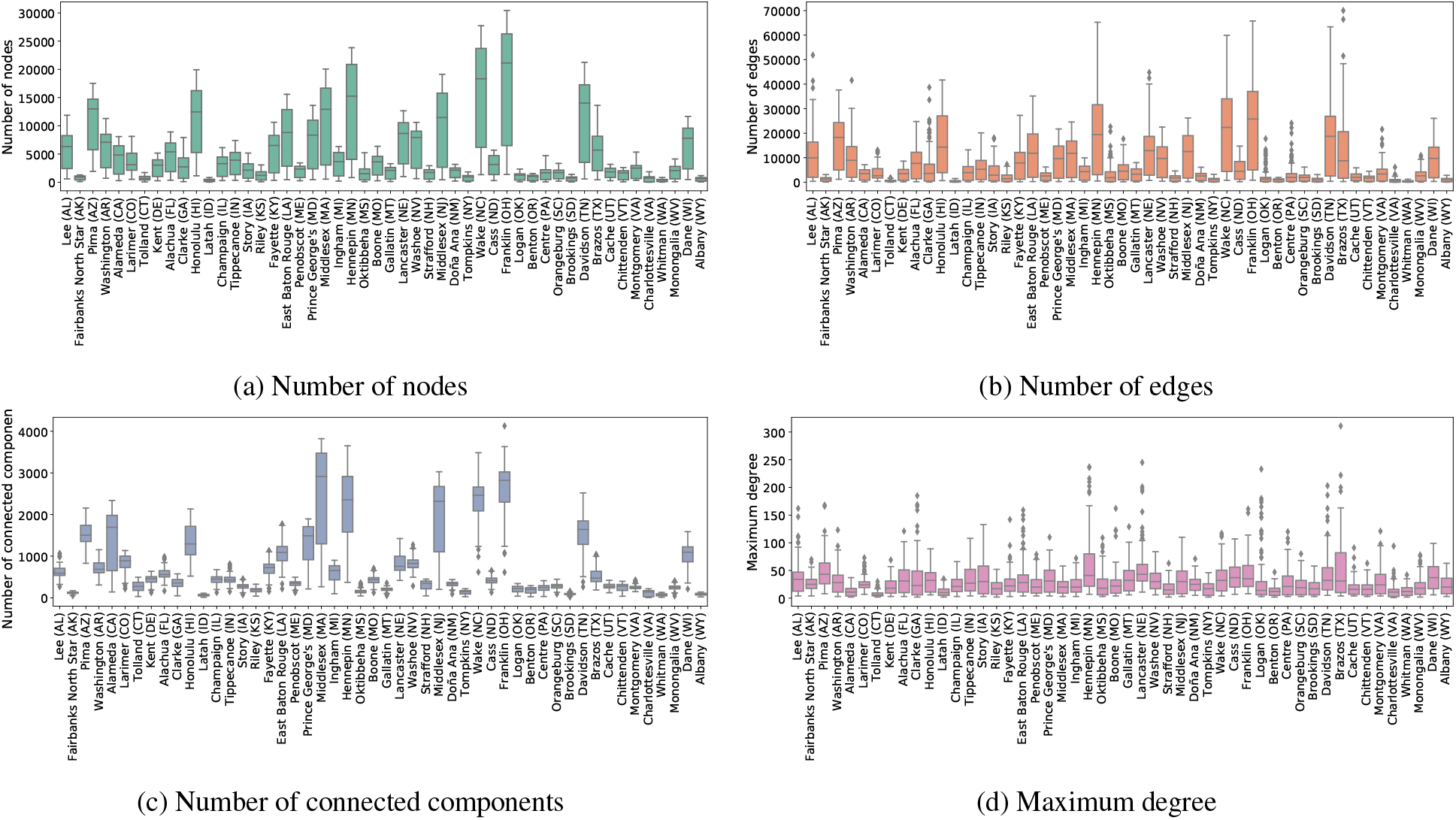
Descriptive statistics about the daily spatial graphs for all the LGUCs.

In Figure 2b, we show the process of generating multiple contact networks from these interactions. In this example, we use *k* = 2, *T*_*c*_ = 1 hour and *T*_*s*_ = 30 minutes. The first graph is obtained for *t* = 08:00 where all the interactions happening between 08:00 and 09:00 are considered and on the right side the corresponding 2-core is shown 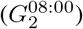. The gray edges correspond to interactions where at least one timestamp does not fall within the specified window. Sliding *t* by *T*_*s*_ = 30 minutes, we similarly obtain another contact network for interactions between 08:30 and 09:30 and the corresponding 2-core 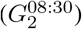.

Referring to the 2-cores, we can see that 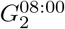 has 2 components of size 3 each and 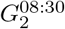 has 1 component of size 4. The largest group size among all is 4 (corresponding to 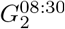), which is the value for metric *k-max-cc-size*. On the other hand, the largest sum of group size is 6 from the two components of 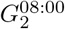, the value for metric *k-max-core-size*. The overall maximum number of gatherings for these contact networks is 2, being the value of the metric *k-max-cc-count*.

The fourth metric is computed by taking *t* = 00:00 and *T*_*c*_ = 1 day, which gives an estimate of the number of people who visit any particular location (spatial window of distance *δ*) during an entire day. The intuition is that, since our data set is a sample, there might be many interactions that are missed. The visit frequency estimate acts as a proxy for those interactions. Taking this into consideration, the fourth metric computes the number of components of the *k*-core of the interaction graph for the entire day. We chose *k* = 1, 2, 3, giving us twelve graph-based metrics for each LGUC. Along with the interaction rate, this gives us thirteen proximity metrics.

### 4.2 The Time-Varying Transmissibility Parameter and the Proximity Metrics

Now we study the relation between the high-resolution proximity metrics and the disease dynamics using the popular disease spread models called the basic compartmental models [3, 28, 25]. In this model, at any given time, individuals in a population of size *N* are assigned to four different compartments, susceptible (*S*), exposed (*E*), infectious (*I*) and removed or recovered (*R*). Assuming homogeneous mixing within the population, the model then specifies how susceptible individuals become exposed, infectious, and then recover. Let *S*(*t*), *E*(*t*), *I*(*t*) and *R*(*t*) denote the number of people who are in the susceptible, exposed, infected, and recovered states at time *t*, respectively. Then the SEIR model is described by the following system of ordinary differential equations.

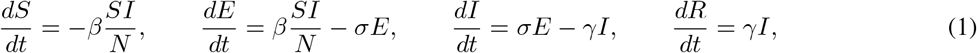

where *β* is referred to as the transmissibility, 1*/σ* the latency period, and 1*/γ* is the recovery rate. *σ* and *γ* are disease-specific parameters and are fixed in our model (1*/σ* = 1*/γ* = 5 days). The transmissibility parameter *β* is allowed to vary over time in our calibration process. This time-varying *β* then encodes all the information about time-varying factors that affect the number of observed cases, such as mobility, mask-wearing, environmental conditions, etc. Hence, (1) is rewritten using a time-varying *β*(*t*) as,

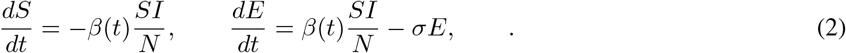

Using a simulation optimization approach, we sequentially estimate *β*(*t*) with appropriate delays (7 days) and scaling (15%) applied from simulated infections to confirmed cases. We train a separate model for each LGUC using the daily new cases as reported by [12] over a period of 227 days ranging from April 1, 2020 to November 14, 2020.

Now, in order to quantify the contribution of mobility to COVID-19 cases, we use multivariate linear regression to determine the fraction of variation in *β*(*t*) explained by the proximity metrics. We have explored other models such as random forest regression and support vector regression also, but observed that a multivariate linear regression (MLR) model was the easiest to train (random forest and support vector regression suffered from overfitting), while providing a reasonable fit. In addition, the MLR model results are easier to interpret.

We consider one MLR model per county where we express the calibrated *β* as a linear combination of the thirteen proximity metrics described in Section 4.1. Since the proximity metrics have different scales, we first normalize each metric by subtracting out its mean and dividing by the its standard deviation. This normalization allows for the resulting model coefficients to be on the same scale. In addition, we compute a 7-day moving average on the metrics. The results of the analysis are presented next.

## 5 Regression Results

In Figure 4a, the MLR coefficients corresponding to each LGUC are presented as a heatmap. The corresponding *p*-values are shown in Figure 4b. The quality of the resulting fit is evaluated using the *R*^2^ score or the coefficient of determination, shown in Figure 5. The median *R*^2^ -score computed across all the LGUCs is ∼0.5, thus indicating a reasonable fit provided by the proximity metrics. Observing the heatmap in Figure 4a, there appears to be no metric that gets significantly high weights across all LGUCs. The MLR coefficients have been obtained as the solution to the ordinary least squares problem. We have explored two sparsity enforcing optimization techniques, LASSO [34] and subset selection methods [27] to determine a sparse set of metrics that yield the best fit. In both cases, the performance in terms of the *R*^2^ score was poorer compared to the ordinary least squares solution and no metric came out as a clear winner across all LGUCs.

**Figure 4:**
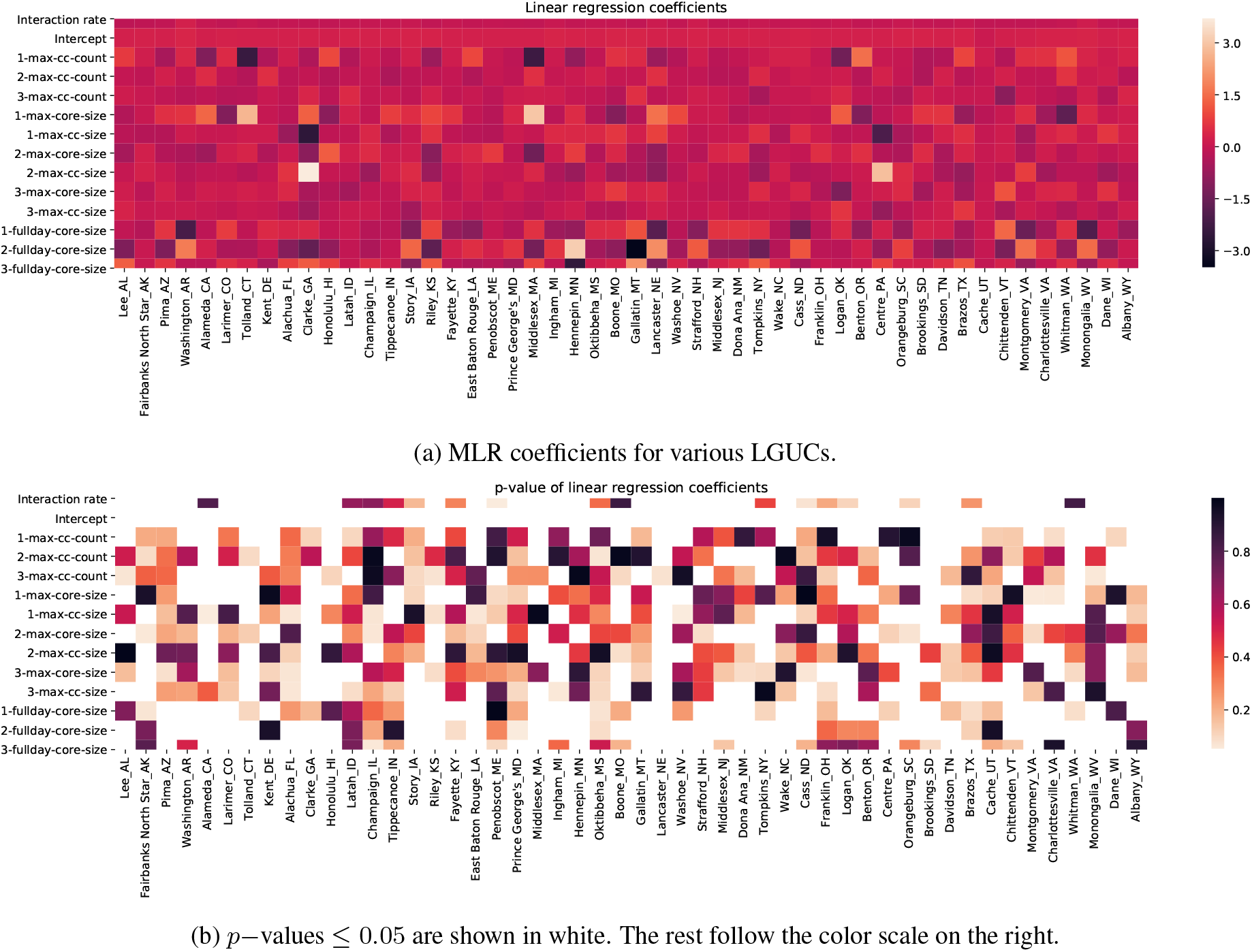
Heatmaps of regression results. In both the figures, we see that there is significant variation in the learned models. This shows that no single proximity metric does well by itself for all the LGUCs.

**Figure 5:**
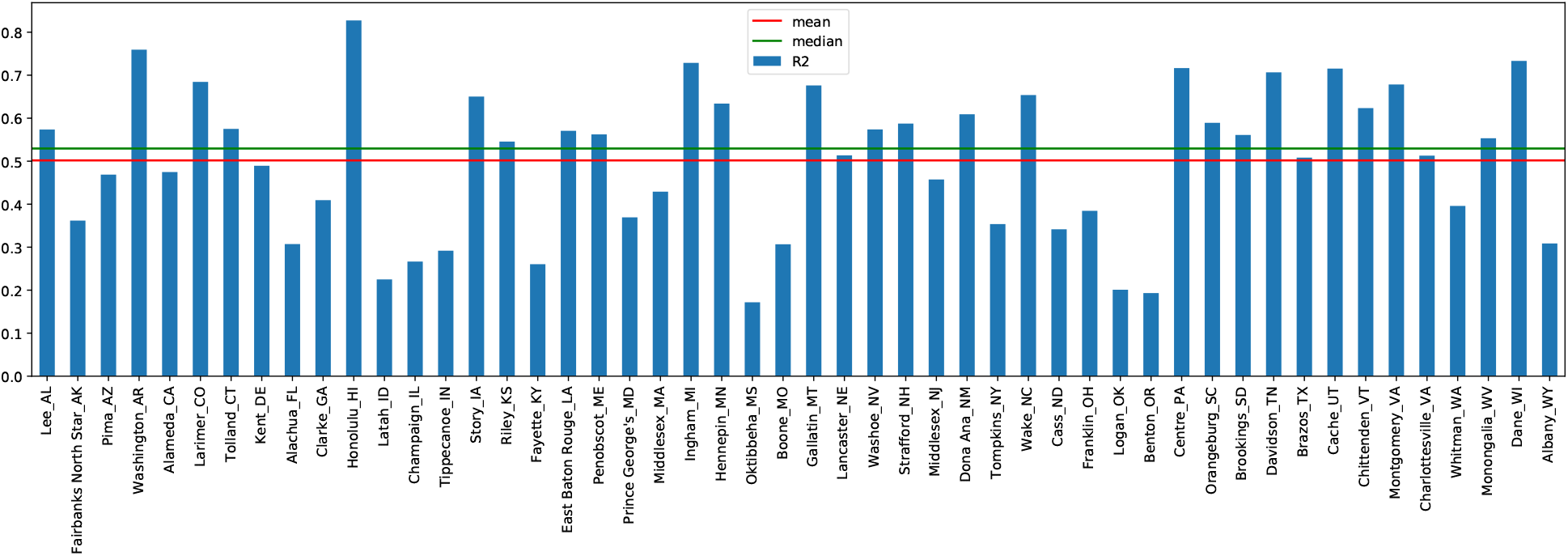
The *R*^2^ score between the fit and the calibrated *β* for each LGUC. We see that the mean and median are quite high, indicating a good fit for at least half the counties.

In Figures 6a and 6b, we show examples of the fit for five LGUCs with the highest and lowest *R*^2^ scores, respectively. The *R*^2^ scores (a score greater than 0.5 for more than 50% of the LGUCs) and visual inspection indicates that a linear combination of the metrics can reasonably capture the variability in the assumed disease transmission parameter or the contact rate. In many of the examples we observe that the peaks in the calibrated *β*(*t*) are being captured by the model. For example, the reconstructed curve for Benton, OR (Figure 6b) shows small peaks aligned with the large peaks in the calibrated *β*(*t*) curve in several cases. However, not all variations in *β*(*t*) can be explained by the metrics, due to the effects of other factors, such as the adherence of the interacting population to mask-wearing or surges in cases due to an increase in testing rates.

**Figure 6:**
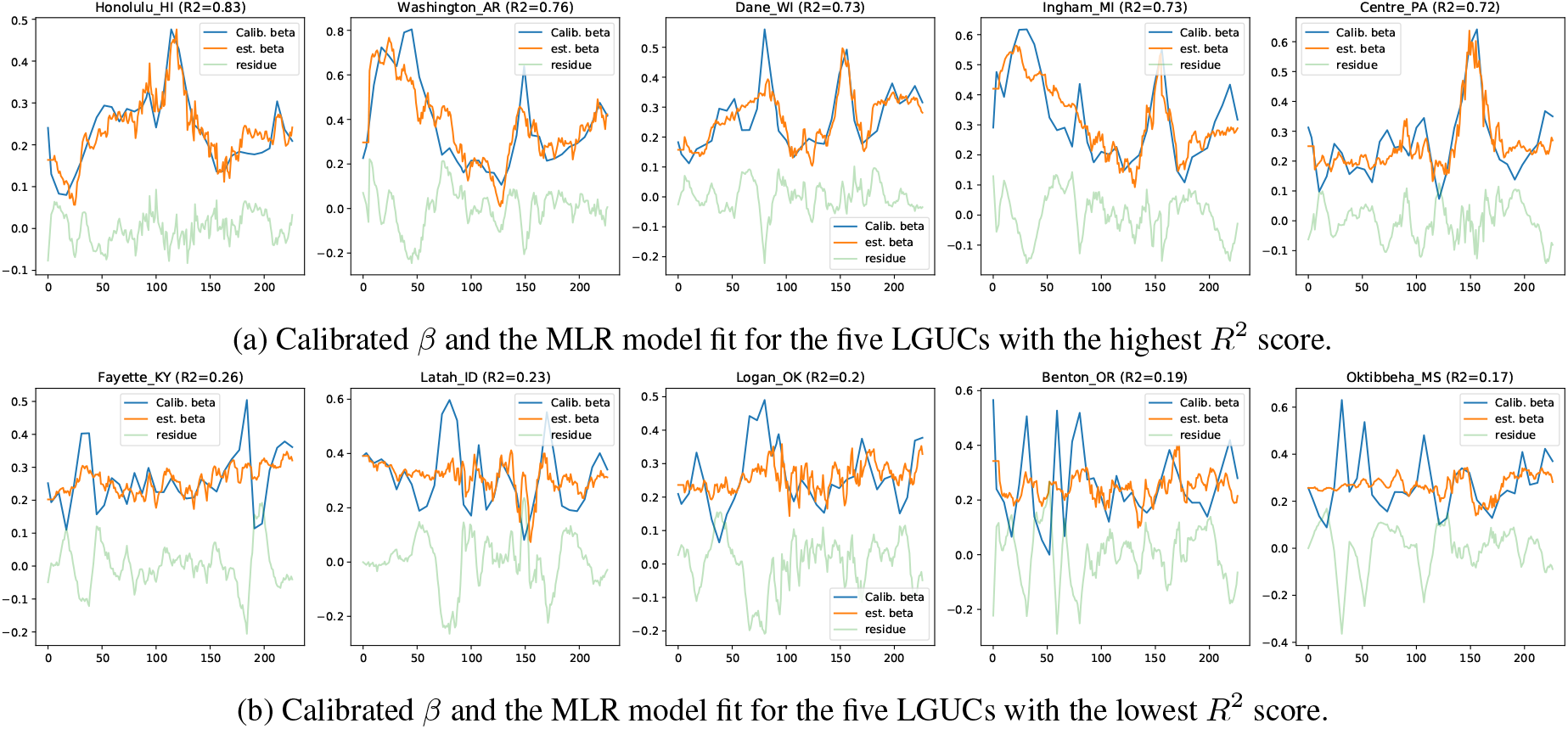
A comparison of MLR model fit to the calibrated *β* along with the residues for the LGUCs with highest and lowest *R*^2^ scores. We see that even for the counties with the lowest *R*^2^ scores, there are peaks in the MLR model fit that often align with the peaks in the calibrated *β*, though they don’t have the same amplitude. This suggests that there is some predictive power in the proximity metrics, but that they don’t account for all the variation in the case rates.

Overall, these results show that there is large variation in the extent to which mobility has contributed to COVID-19 cases. We now do a more detailed analysis of the correlation between the proximity metrics and case rates.

## 6 Correlation Results

As described earlier, we calculated a total of 13 different proximity metrics. For each of the four graph metrics described in Table 2, we choose *k* = 1, 2, 3. We calculate these proximity metrics for all the 50 LGUCs from April 2020 to October 2020, as the state-mandated social distancing orders had been placed by that time and the Fall semester reopening phase also takes place within this timeframe. To measure correlation between these metrics and the case rates of the LGUCs, we calculate the Pearson Correlation Coefficient (PCC). County level case counts were converted to case rates per 100,000 to normalize across different population sizes and the data for both case rate and the proximity metric were smoothed by computing 7-day moving averages to eliminate any within-week reporting artifacts and noise in the data. PCC between the case rates and each time series proximity metric is calculated for different lead times and the maximum PCC was recorded for each county and each different metric. Figure 7 shows the maximum PCC obtained over all the lags between the case rate and each proximity metric for all the LGUCs. A high positive value of maximum PCC means that the metric and the case rate are highly correlated. From the figure, it is obvious that no single metric is a universally good correlative indicator of case rate across all LGUCs. We also show the correlation results in a map for each of the COI in Figure 8. For each COI, we show the proximity metric that gives the best result in terms of correlation. The color of the circle represents the population of the LGUC. The circle size represents how good the correlation is on a scale from 0 to 1. The ones having no or irrelevant correlation are shown with an ‘X’. Interestingly, we observe that these LGUCs are mostly in the Northeastern region. These are also the areas which were some of the hardest hit in the early stages of the COVID-19 pandemic, and instituted strong mask-wearing rules.

**Figure 7:**
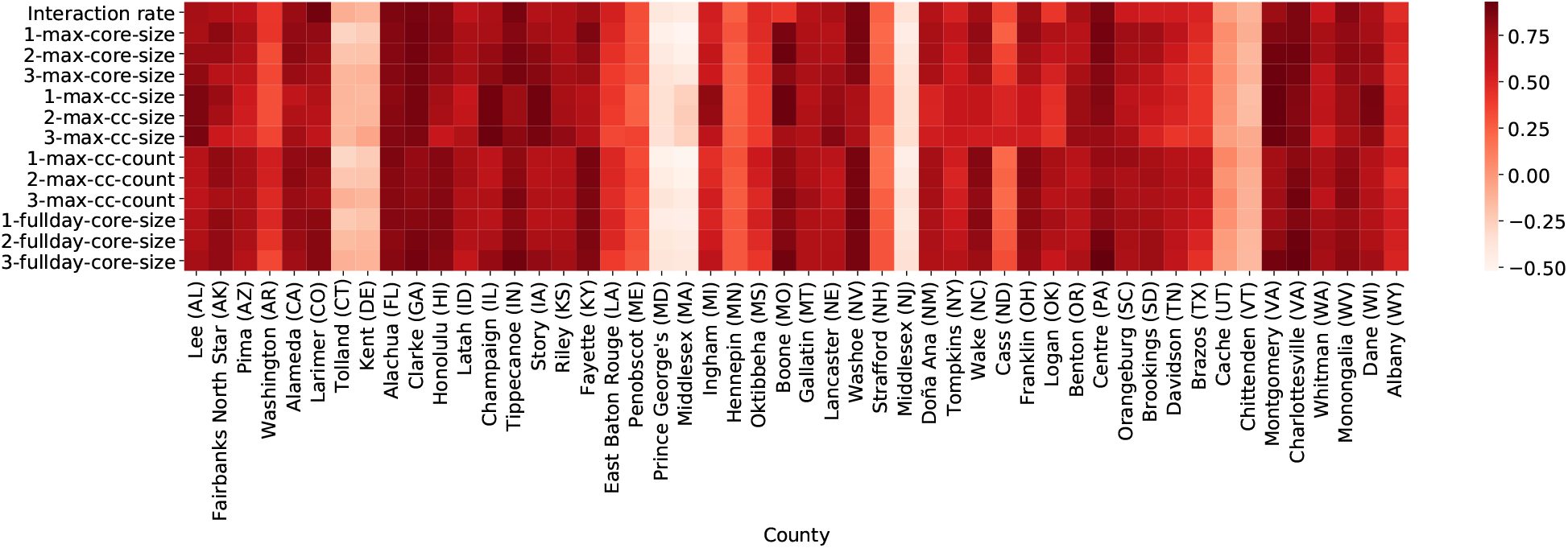
Maximum correlation of each proximity metric for the LGUCs. We see that different proximity metrics have the highest correlation with the case rate in different counties, and that there are a few where we do not get a good correlation at all.

**Figure 8:**
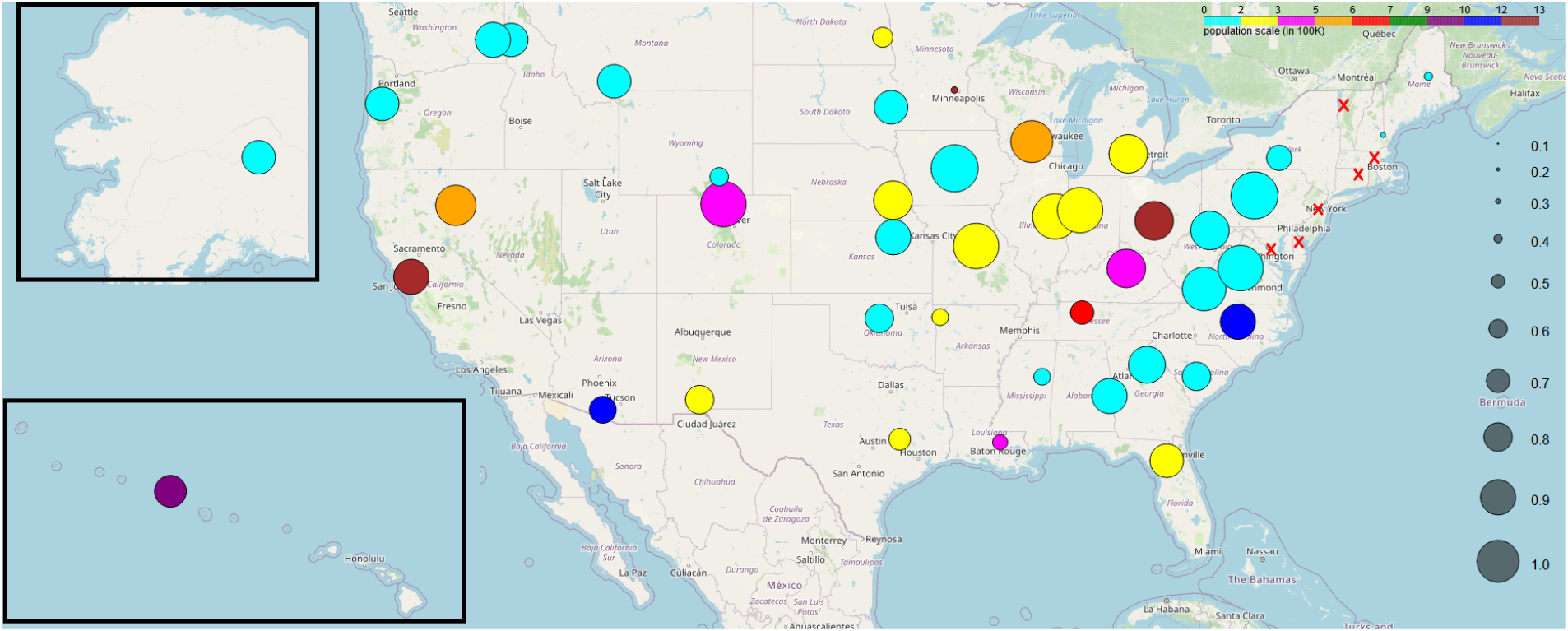
The best proximity metric for each LGUC on a US map. Circle color represents the county population and circle size represents the value of the metric with the best correlation. We see that the LGUCs where the proximity metrics have poor correlation with the case rates are concentrated in the Northeast. This may be due to the early impact of the epidemic in that area and the strong NPIs that were enforced in response.

Since no single metric seems to be the best in terms of correlation with cases, and given that the reconstructions in Figure 6 show peaks that align well with the peaks in the calibrated *β*(*t*) even for cases with low *R*^2^, in the next section we consider using the combination of metrics to predict peaks in the case rates.

## 7 COVID Spike Prediction

Instead of predicting the exact case rates, we focus on studying whether the proximity metrics are suited for a more coarse-grained task such as predicting spikes in the case rate. If a spike in the case rate is preceded by a spike in one or more of these proximity metrics, it is possible to build a model that can predict whether there is a spike in the near future, using the proximity metrics as features. More specifically, we want to address the question, *using the proximity metrics for the previous p days, is it possible to predict whether there will be a spike in cases in the next L days?*

In order to build a model that can answer this question, we use the parameters *p* and *L* to construct our dataset. More specifically, if the current day is 𝒟, we use the proximity metrics of the previous *p* days from day 𝒟, i.e., the metrics for days 𝒟, 𝒟 − 1, …, 𝒟 − *p* + 1, as features. The corresponding label is 1 if there is a spike in the COVID cases between day 𝒟 + 1 to 𝒟 + *L* − 1, otherwise 0. We also include additional demographic features specific to a COI which we describe in the next section. To observe the correlation and predict spikes in COVID cases, we used COVID-19 case counts obtained from USAFacts [36]. We also use county population data [36], RUCC codes (urban-rural designations of counties) from USDA [14], median income [13] and expected age [35] as county demographics.

In a time series data, we consider a value to be a spike if it exceeds a certain threshold compared to its neighboring values. More formally if the value at time *T* is greater *y*, where *y* is inferred from the *x* neighboring values at time range [*T − x, T*) and (*T, T* + *x*]. In order to detect spikes from the case rate data we used the *signal*.*find_peaks* algorithm from the Python scipy [37] library. The algorithm needs two parameters, the range of neighboring values *x* and the height threshold *y*. We use the average plus two standard deviations of the neighboring case rate values as the height threshold and 14 days for the range of neighboring values. Figure 9 shows some of the spikes detected by the algorithm. We use these spikes as the labels for the spike prediction learning problem.

**Figure 9:**
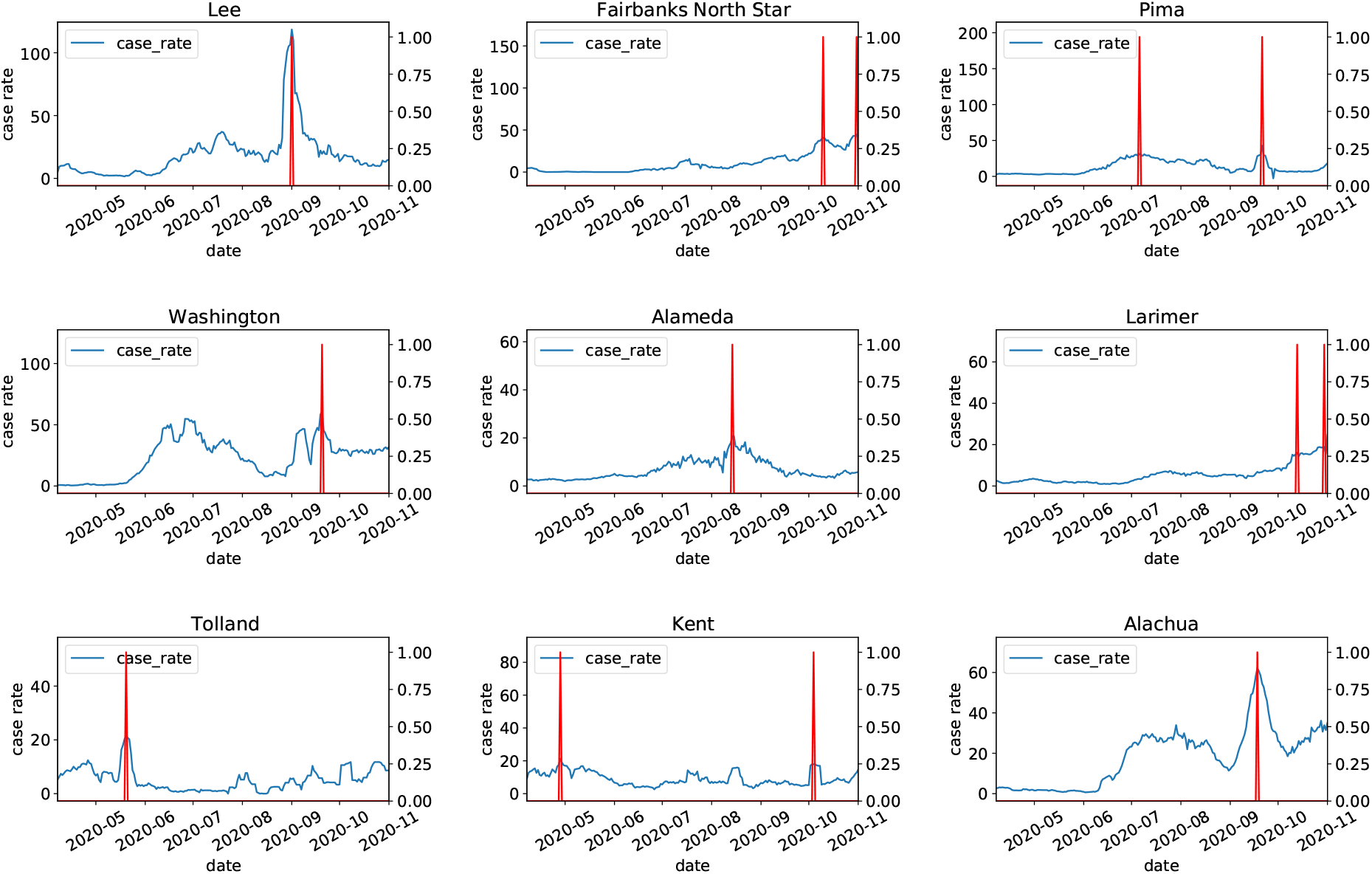
Spikes in case rates detected by the *signal*.*find_peaks* algorithm. Blue curves are the case rate and the red spikes show places where the algorithm has detected a spike. These are used as labels in the spike prediction learning problem.

In our experiment, we choose *p* = 7 days and *L* = 21 days, i.e., we use the proximity metrics over 7 days to predict if there will be a spike in cases at any time during the subsequent 21 days. Since we have 13 proximity metrics we get a total of 91 features for a week of data. These 91 values are the input features for one data point in our dataset. The output is set to 1 if there is a spike at any time during the subsequent 21 days, otherwise it is set to 0. In our final dataset, we also use the 4 additional features described above as demographic features specific to each LGUC, along with the 91 proximity features. These 4 features are the same across all the data points for an LGUC. The intuition is to capture the intrinisic differences between counties through these demographic features. These features are denoted as follows in the results below: county population (D_0), RUCC type (D_1), median income of the county population (D_2), and expected age of the population of the county (D_3).

### 7.1 Spike Prediction

The features are normalized before they are used to train a model. We compared multiple learning methods. Table 3 shows the performance of different models on the test dataset for a 10-fold cross validation experiment. The best performances are observed from XGBoost, k-nearest neighbor, and random forest. We can observe that although most of the models generate high accuracy score, these 3 models are the only ones generating a reasonable F1 score of above 80%. For the other learning methods, the F1 score drops sharply, indicating that these models either suffer from a class imbalance problem or they do not learn the parameters correctly. In our scenario, F1 scores are important because there are fewer positive (spike) examples than negative (not spike) ones.

**Table 3:**
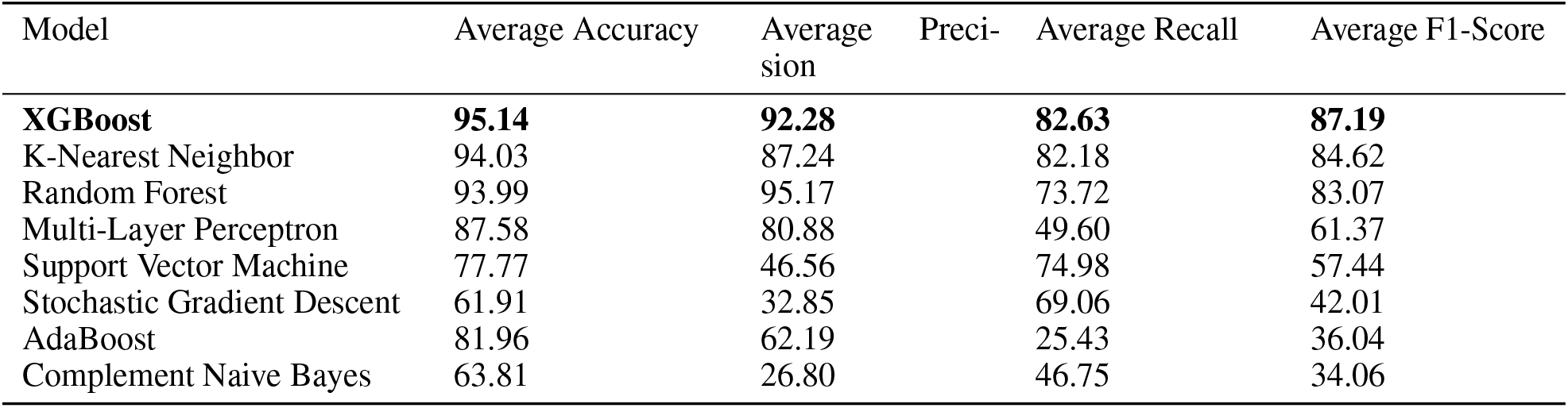
Performance of models in cross validation test.

#### 7.1.1 Ablation Study

Since XGBoost provides the best overall performance (from Table 3), we performed an ablation study using XGBoost to study how different combination of features affect the performance of the model. Figure 10 shows the result of the ablation studies. The top teal bar shows the performance when all the time series metrics and demographic features were used. In this case, the number of features were 95.

**Figure 10:**
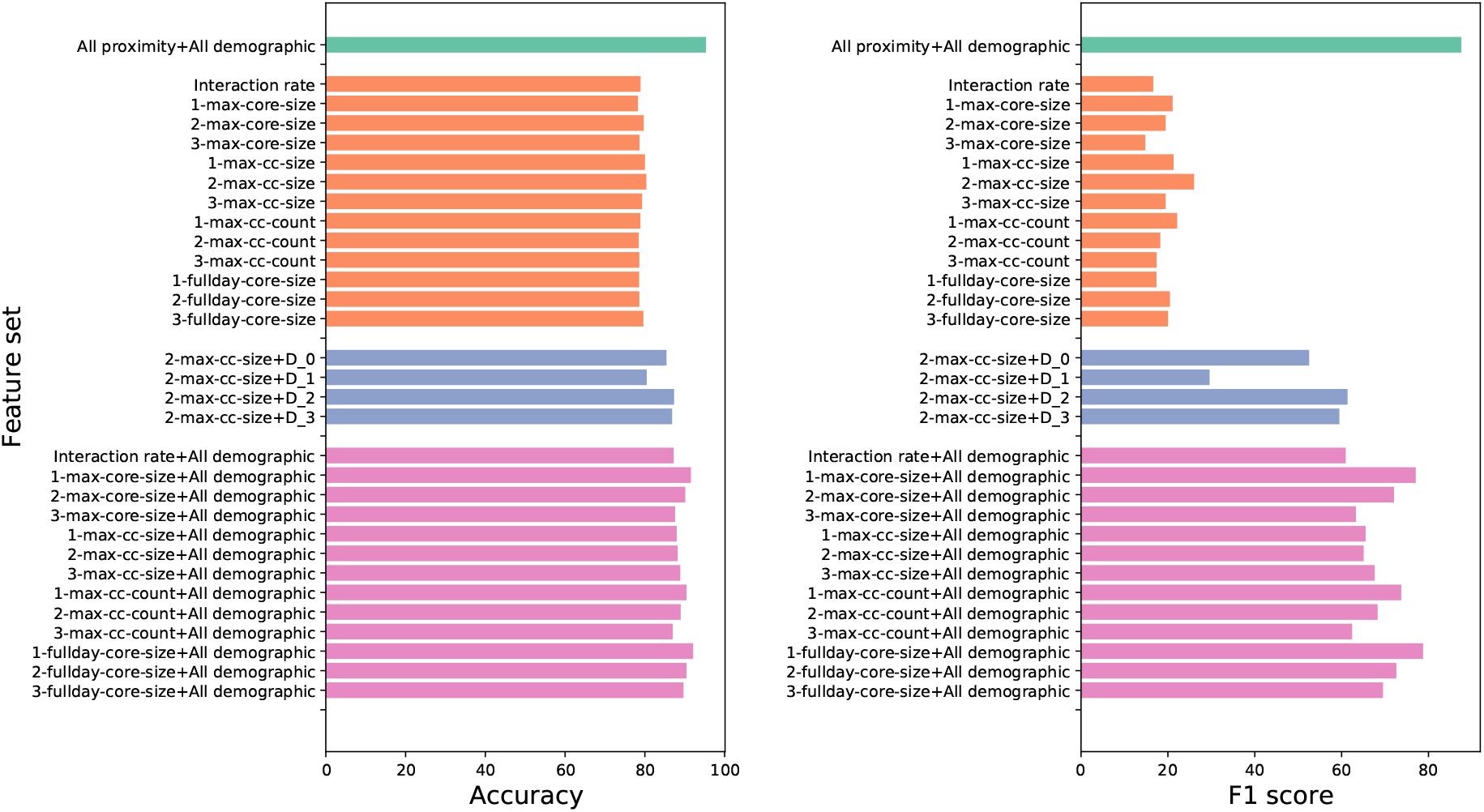
Ablation study result shows accuracy and F1 score for different sets of proximity metrics. We see that the F1 score drops sharply when proximity metrics are considered individually, even if all the demographic features are added. Combining all the proximity and demographic metrics is significantly better than the other results.

The next set of orange bars shows the performance of the model for each time series proximity metric by itself. We can see that the performance drops significantly from the low F1 scores. Therefore, it is evident that all the features provide useful information about future spikes in the case rate and using them altogether is better than using any single feature.

In the next set of experiments, we use each demographic feature with the time series proximity metric that performed the best in the second set of ablation studies (*2-max-cc-size*). We can see that although the accuracy did not improve by much over the accuracy when only *2-max-cc-size* was used, the F1 score improved substantially. This means that using both time series and demographic features is more beneficial than using time series features alone.

Finally, we used all the demographic features with each of the time series features. We can see that the F1 scores improved by a lot compared to the second set of experiments. This again validates the usefulness of the demographic features. However, none of these experiments gave a performance similar to or better than when all the time series and demographic features were used. Therefore, it is validated that all the features we used in our experiment were necessary for predicting case rate spikes.

#### 7.1.2 Leave-one county out experiment

The last study we do is to determine how well our model is able to predict spikes on a completely unknown county. For this, we leave the data corresponding to one LGUC completely out while building the training data set and use these left out data to build the test dataset. This is done once for each LGUC and the result is shown in Figure 11. We can observe that although the model shows good accuracy while predicting on an entirely unknown county, it does not do very well in terms of recall. A high recall would mean that the model can predict most of the spikes accurately which is desirable in the case. However, in around a fifth of the counties, the model performs quite well.

**Figure 11:**
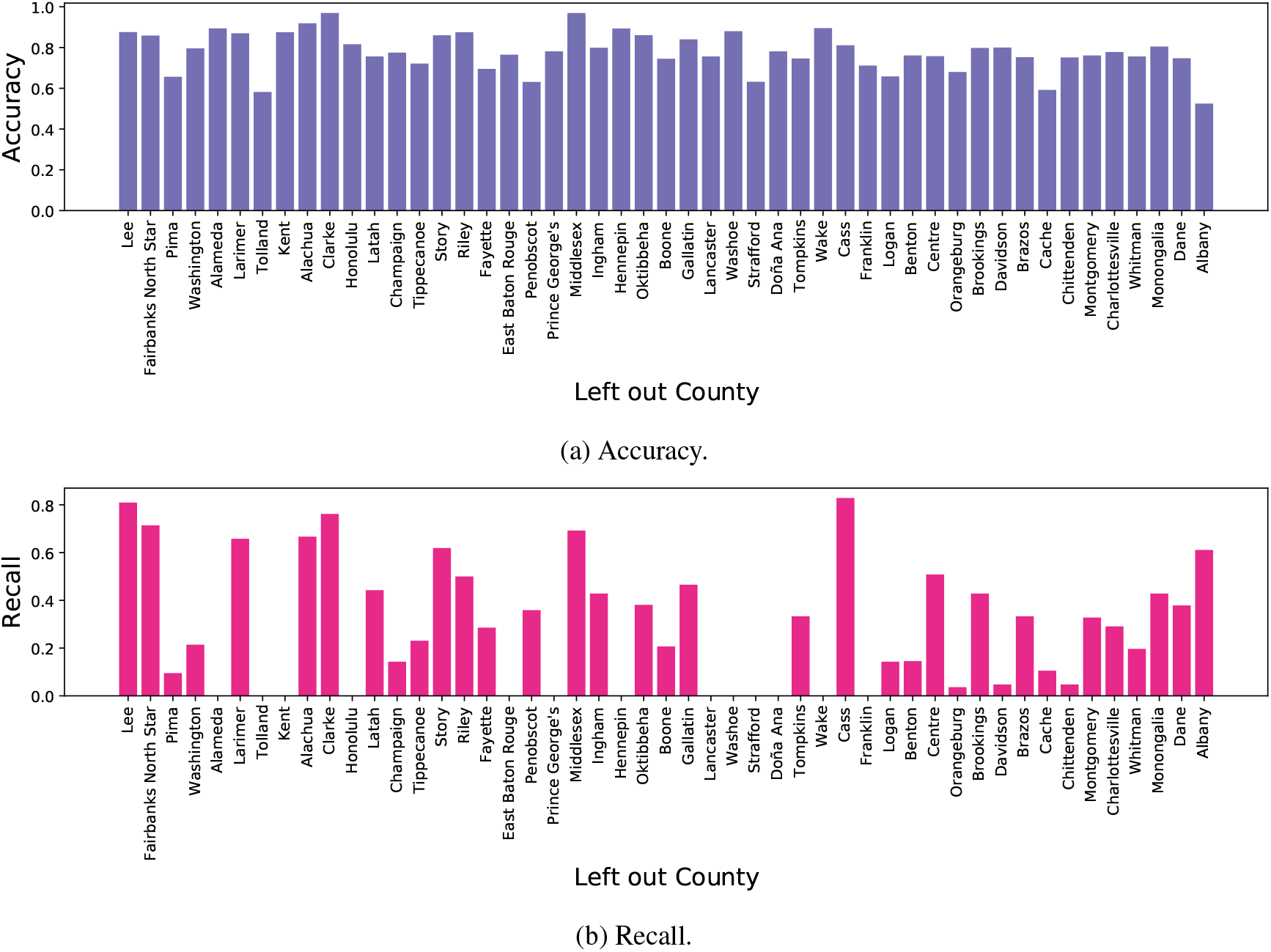
Accuracy and recall scores for the leave-one-county-out experiment. While the accuracy remains fairly high, the recall performance is poor for several counties. This shows that there are several counties which are dissimilar to the other counties, but where proximity metrics (from the same county) are still useful, based on the previous results in Table 3.

Our intuition is that there are sets of counties that show similar behavior in their case rate trend and the trends in different proximity metrics. When one of the counties from these sets are left out, the knowledge gathered from training using the other counties becomes helpful. These similar counties may not be geographically near each other. What specific properties make these counties similar may be an interesting research question and a possible future direction.

## 8 Discussion

A general understanding of the role of mobility in epidemics is still to be developed. At a mechanistic level, changes in mobility cause changes in the structure of the social contact network, which mediates the spread of contagions. As populations reduce their mobility, the social contact network gets sparser. If the herd immunity threshold is reached in this sparser network, cases start to fall. However, if mobility starts to increase again before the epidemic is fully eradicated, cases can increase again. This phenomenon manifests as “waves” of the epidemic, as we saw in the summer and fall of 2020.

However, there are several complicating factors. The above general mechanism is modulated by the prevalence rate, so that reducing mobility may not hit the herd immunity threshold if the prevalence is high enough already. It is possible that this is why mobility restrictions in the fall of 2020 were not as effective as they were in the summer of 2020. As we have discussed earlier, several other factors, such as mask-wearing and environmental factors also modulate the relationship between mobility and cases.

It is thus important to understand just how much responsibility can be attributed to mobility for the case rates. This is what we have attempted to quantify in this work. While most prior work in the literature has tried to bypass this issue and go straight to forecasting, we believe that this kind of understanding is important to have a scientific basis for policy-making. It also gives pointers for a further comparative study. What is the difference between counties where mobility has played a large role in determining cases and the counties where it hasn’t? The same methodology that we have followed in this paper could be used to cast light on this issue, for example by bringing in data sets about mask-wearing.

A point of caution here is that there is probably some interaction between factors like mobility and mask-wearing, in that it is possible that people who wear masks regularly may feel more confident in interacting with others and may do so more frequently. On the other hand, both these variables are also influenced by external factors like political messaging and political orientations.

We have also shown that multiple graph-based metrics are useful in predicting the effects of mobility on cases. This further points to the benefits of network-based analysis of epidemics. It is of course possible that other metrics (graph-based or not) could be used to improve our results. Therefore, our results should be seen as a lower bound on the role of mobility in the COVID-19 epidemic. Other mobility data and other metrics could show that the role of mobility is even higher.

The overall high classification scores in the peak-prediction experiment indicate that, even where the *R*^2^ values were small, mobility data are useful for making actionable predictions. Further, the final experiment, where we did the leave-one-county-out test, indicates that we might be able to cluster similar counties in terms of the role mobility is playing in the epidemic spread in those counties. This could lead to further insights from comparative studies.

## Data Availability

Aggregate proximity metric statistics used in this paper are available from the authors upon request.

## 9 Acknowledgments

This work was partially supported by National Institutes of Health (NIH) Grant R01GM109718, NSF BIG DATA Grant IIS-1633028, NSF Grant No OAC-1916805, NSF Expeditions in Computing Grant CCF-1918656, CCF-1917819, NSF RAPID CNS-2028004, NSF RAPID OAC-2027541, US Centers for Disease Control and Prevention 75D30119C05935, DTRA subcontract-ARA S-D00189-15-TO-01-UVA, a grant from Google and VDH contract VDH-21-501-0141. Any opinions, findings, and conclusions or recommendations expressed in this material are those of the author(s) and do not necessarily reflect the views of the funding agencies.

## Notes

### Competing Interest Statement

The authors have declared no competing interest.

### Author Declarations

The study at University of Virginia was approved by the Institutional Review Board for Social and Behavioral Sciences as part of the Human Research Protection Program.

